# Epidemic transmission with quarantine measures: application to COVID-19

**DOI:** 10.1101/2021.02.09.21251288

**Authors:** S.A. Trigger, E.B. Czerniawski, A.M. Ignatov

## Abstract

Equations for infection spread in a closed population are found in discrete approximation, corresponding to the published statistical data, and in continuous time in the form of delay differential equations. We consider the epidemic as dependent upon four key parameters: the size of population involved, the mean number of dangerous contacts of one infected person per day, the probability to transmit infection due to such contact and the mean duration of disease. In the simplest case of free-running epidemic in an infinite population, the number of infected rises exponentially day by day. Here we show the model for epidemic process in a closed population, constrained by isolation, treatment and so on. The four parameters introduced here have the clear sense and are in association with the well-known concept of reproduction number in the continuous susceptible– infectious–removed, susceptible–exposed–infectious–removed (SIR, SEIR) models. We derive the initial rate of infection spread from the published statistical data for the initial stage of epidemic, when the quarantine measures were absent. On this basis, we can found the corresponding basic reproduction number mentioned above. Our approach allows evaluating the influence of quarantine measures on free pandemic process that leads to the time-dependent rate of infection and suppression of infection. We found a good correspondence of the theory and reliable statistical data. The initially formulated discrete model, describing epidemic course day by day is transferred to differential form. The conditions for saturation of epidemic are found by solving the delay differential equations. They differ essentially from ones in SIR model due to finite delay, typical for COVID-19 The proposed model opens up the possibility to predict the optimal level of social quarantine measures. The model is quite flexible and it can be extended to more complex cases.

## I. INTRODUCTION

The appearance of a new virus in China at the end of 2019 has been recognized by the World Health Organization (WHO) as a pandemic in March 11, 2020. This virus named SARS-CoV-2 (causing COVID-19 disease) to the moment infected 25 millions of people and almost 1 million of deaths over the world. High mortality required introduction of essential quarantine measures to confront pandemic and to reduce victims. The strong quarantine measures lead to the serious damage to the global economy.

The most of existing models for the spread of infection simulate the spontaneous development of an epidemic and describe all its stages. There are two kinds of such models: susceptible-infected-susceptible (SIS) models and susceptible–infectious–removed, susceptible– exposed–infectious–removed (SIR, SEIR) models. The first go back to the pioneering work of Kermack and McKendrick [1] and uses the assumption that the recovered people can immediately get infection again. On the contrary, the latter are built on the assumption that the recovered people save strong immunity during epidemic (see, e.g. [2]). There are many variants of those models.The SIS models are used in the mathematical epidemiology [3]. An overview is given in [4–6] (see also references therein). Balance between the susceptible and infected members of population under the various conditions of infection transfer, are the subject of research in [7–9].

The susceptible–infectious–removed, susceptible– exposed–infectious–removed (SIR, SEIR) models (see [10–12] and references therein), as well as a few papers the changing in time parameters ([13, 14] and references therein), which assume the overall immunity of recovered people, are somewhat closer to the formulation of our model presented here. However, opportunity of immediate recovery contained in SIR-type models is quite questionable for diseases like COVID-19. With regard to the SIR and SEIR models, it should be noted that the time derivative of the infectious people *dI*/*dt* is determined, in particular, by the term *γI* (where *γ*^−1^ is the average duration of a disease). Therefore, there is a possibility of immediate recovering, which contradicts the observed features of COVID-19.

There are stochastic (e.g., [15, 16]) and deterministic approaches to the description of epidemic course. The latter include the SIR, SEIR models and their modifications. Therefore, it is natural to develop the existing deterministic models such as [17] to more precise and appropriate description of the COVID-19. This development should take into account such a feature of COVID-19 that infected is contagious for a long time. The advantage of this type of model is the ability to compare their results with the statistics reported by the authorities of countries and regions [13].

We consider some of specific features of COVID-19 [18– 22] when developing the discrete model of epidemic [23]. In contrast with the SIR-type models, the model under consideration for a closed population includes two independent parameters. One of them is the average duration of disease *d* and the second, the parameter of the infection transmission rate *p*, similar to the reproduction number *R*_0_ in the SIR models. We divide the parameter *p* on two *n*_*c*_*k*, where *n*_*c*_ is the average quantity of dangerous contacts per day for one infected person and *k* is the average susceptibility to virus of a healthy person. The initial condition, namely the quantity of the infected people at the beginning of the epidemic process in the concrete closed region, is also essential. Here we restrict ourselves to closed populations (country, region, city, and etc.). Of course, there is a steady exchange between considering populations. However, already at an early stage of the epidemic, the authorities use isolation measures to reduce such flows to a minimum. The transboundary transmission of the infection before the quarantine cannot be accurately calculated. This first stage of epidemic can be approximated as the free running epidemic, when the quarantine measures are practically absent. Analysis of the impact of the flow of people between regions on the epidemic is a separate task that can be considered, including on the basis of the proposed model.

Thus, the national and local authorities are taking actions to slow down the epidemic. We can define these measures as an epidemic under partial control. It is very important to slow down the epidemic rate to give physicians the ability to provide patients with the necessary amount of medical care. Therefore, to study the entire course of infection spread from the very beginning may be useful.

We believe that it is important to take into account such features of the disease caused by COVID-19, as the frequent recurrence of asymptomatic course of the disease, long duration, high infectivity and high mortality. The model we present here makes it possible to take into account all features just mentioned. We consider the course of epidemic in the closed population. Based on this, the authorities can outline a strategy for quarantine measures. It should be noted that the real epidemic develops under the significant influence of some external limiting factors that are not considered in this paper, but the model, according to its capabilities, allows, if necessary, to take them into account. The present work has been announced recently [24].

### II. EPIDEMIC IN UNLIMITED AND LIMITED POPULATIONS

Following Ref. [16], first we will examine an unlimited population where anyone can be infected since no one has immunity. Let us assume that the average duration of the disease (*d*, days) is the time when the carrier of the infection is able to transmit it to others. In fact this time can vary from place to place depending on the accepted accounting methods, on the regional type of age pyramids etc. (see e.g.[25, 26]). Moreover, the reliability of the COVID-19 tests till now is about 50 percent. Therefore, we determined the value of *d* empirically on the basis of published statistical data for different countries, e.g. 18 days for Switzerland and 16 days for Israel. So by day *l* ≤ *d* after the first infected appears, the *N*_*I*_ (*n*_*c*_; *k*; *l*) people will be infected

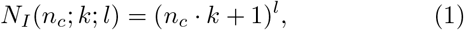

where *n*_*c*_ is the average number of the dangerous (i.e. close unprotected contacts that may lead to infection) contacts, that may lead to infection, of one infected person per day, and *k* is the probability of infection during these dangerous contacts. Thus, *p* = *n*_*c*_ *k* is the average number of people infected by one virulent person per day in an unlimited population. The general case of the initial stage of epidemic for arbitrary value *p* is considered in [22]. It’s obvious that the number of infected is growing exponentially like *l* ln(*p* + 1).

However, after *l* ≥ *d* days the situation changes, because removed people (recovered and dead) appear in the population. So, the number of infected becomes equal

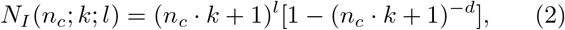

With an average duration of disease about 14 days, equations (1) and (2) for *l* ≥ *d* give approximately the same results.

Now, let us consider an epidemic in a closed population number of *N* people.

After more than *d* days after the epidemic outbreak the population *P* splits into the following subsets: *H* - a subset of healthy people (“preys”); *V* - a subset of infectious people (virus carriers, “predators”); *R* - a subset of removed people that can neither infect nor be infected; *A* - a subset of people ever infected in the course of epidemic (affected). In daily statistics, subsets *V* and *A* are denoted as “actual cases” and “total cases” respectively. During an epidemic, the number of people from subset *A* (people *V* together with people *R*) will inevitably increase. People *V* do not infect themselves or people *R*, therefore both come out of the number of potential preys.Since people *V* do not infect themselves or people *R*, they come out of the number of potential preys. The space for new infections slightly narrows. Therefore, the current number of dangerous contacts becomes less than the initial value of the dangerous contacts *n*_*c*_. This means that the transmission rate of the infection is determined by the variable 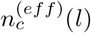, depending on the number of days after the first infection, reads:

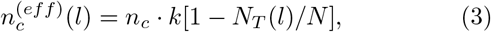

In fact, the dependence 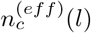 on time appears here as the dependence on the ratio of the total number of affected (sum of infected, recovered and dead) people on the day before the current day *N*_*T*_ (*l* − 1), to the full population size *N*. Therefore, following to the reliable and accepted statistics [19] we use below only two main characteristic functions for the COVID-19 epidemic: *N*_*T*_ (*l*) the total number of affected to the day *l* people and *N*_*I*_ (*l*) -people capable infect. The latter consist of evidently sick, the people in incubation period and asymptomatic ones. These groups can be considered separately as in SEIR models [12–14], [17], see also [22]. It is also possible in the model under consideration. However, such a separation seems non-adequate to the available statistical data of WHO and national authorities [19]. The reported incubation periods can vary between (0 ÷ 27) days [19], that is too much to compare the theory which includes incubation separately with the reliable statistics.

At the beginning of the epidemic (*l* ≤ *d*) all people infected up to the day *l* by definition are the virus carriers *N*_*T*_ (*l*) = *N*_*I*_ (*l*), and we obtain the equation:

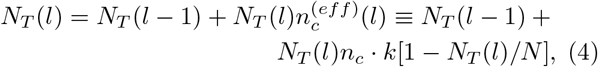

where *N*_*T*_ (*l*) is the number of people infected up to the day *l*. To shorten the notation, we will further omit the arguments *n*_*c*_ and *k* and instead *N*_*I*_ (*n*_*c*_; *k*; *l*) we will write *N*_*I*_ (*l*). After *d* days in the population *P* begin to appear first people coming out of epidemic. They form the subset of removed people denoted above as *R*.

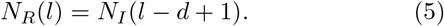

Wherein, for *l* > *d*, the numbers of infected people *N*_*I*_ (*l*) (virus carriers) and the total number of infected cases *N*_*T*_ (*l*) to the day *l* can be described by the main system of equations without quarantine measures [23, 24]

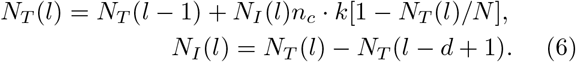

Therefore, substitution *N*_*I*_ from the second equation (6) to the first one leads to the closed equation for *N*_*T*_ (*l*)

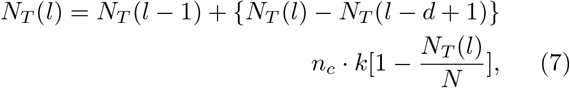

As is easy to show equation (7) not always lead to lim_*l*→∞_ *N*_*T*_ (*l*)/*N* → 1. For some parameters equation (7) can contain solutions with *N*_*I*_ (*l* →∞) = *N*_*sat*_ < *N*. Therefore, the growth of total cases of disease in course of the free-running epidemic can be limited by the level of saturation *N*_*sat*_, called collective immunity (CI).

Figure 1 shows the curves corresponding to the Eqs. (6), (7). For calculations we used the initial condition *N*_*I*_ (*l* = 1) = *N*_*T*_ (*l* = 1) = 1 + *n*_*c*_*k*. As mentioned above the parameters *n*_*c*_ and *k* enter in the above equations only as a product. This pair of numbers could be replaced by their product *p*, but it makes sense to distinguish between them, since *n*_*c*_ number reflects the characteristics of local conditions and quarantine policy for the population *P*, while *k* is associated with the level of personal susceptibility among the people of *P*.

**Figure 1:**
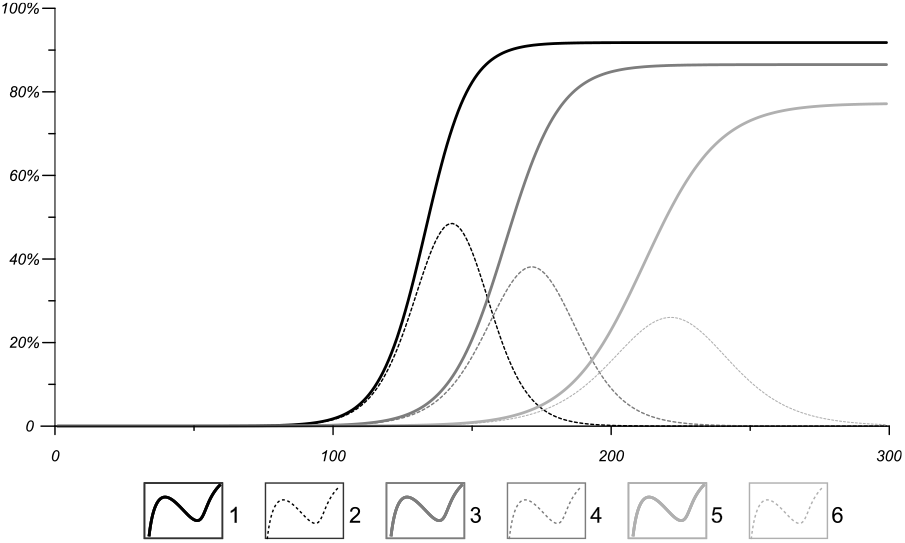
The total number of infections (thick curves) and the number of actually infected (thin curves) computed with Eqs. (6) and (7) during the free-running epidemic in a closed populations a percentage of the population size. Here *p* = 0.14 (curves 1, 2), *p* = 0.12 (curves 3, 4), *p* = 0.1 (curves 5, 6); *d* = 20.

### III. EPIDEMIC IN A LIMITED POPULATION WITH QUARANTINE MEASURES

Let us consider now the influence of quarantine measures on epidemic process. The parameter *n*_*c*_, initially possessing a certain specific characteristic value, can be modified *n*_*c*_ → *n*(*l*) (*p* → *p*(*l*)) by the introduction of quarantine or protective measures. Such measures can be taken into account in the developed theory by introducing a function *n*(*l*) in the course of the epidemic. This “function of an external influence” is in general different for different populations.

To prove this we consider the epidemic course with changing day after day *n*_*c*_ → *n*(*l*) ≡ *n*_*l*_. As is easy to seen, if the epidemic starts with one infected person, after the first day the full number of infected must be *N*_*I*_ (1) = 1 + *n*_1_*k*, for the second day - *N*_*I*_ (1) = 1 + *n*_1_*k* + (1 + *n*_1_*k*)*n*_2_*k* = (1 + *n*_1_*k*)(1 + *n*_2_*k*) and so on. From here we can determine the full number *N*_*I*_ (*l*) of all infected since day 1 till day *l* [23]:

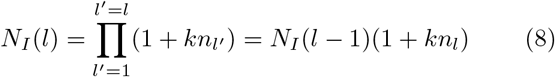

Taking into account that virus carriers cannot infect the infected people and removed ones, we find the true expression *N*_*I*_ (*l*) (it is also describes the number of real virus carriers, named usually as actual cases, for *l* < *d*)

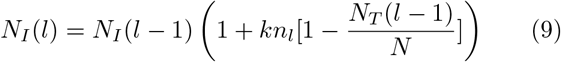

Then the equations for total cases and active cases (current virus carriers) at arbitrary day *l* ≥ *d* for the duration of the disease *d* are

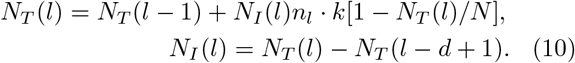

Here *n*_*l*_ is an arbitrary function of *l*, which describing the introduction of the quarantine measures by external regulation. Equations (9) and (10), being a generalization of equation (6), are *the main equations for epidemic process with quarantine measures*.

Figure 2 shows the computed and statistical data for *p*(*l*) for Israel. Up to *l* = 33 days there is a free-running stochastic epidemic process we approximate with the constant *p*_0_ = 0.26. Between 34 < *l* < 83 days statistics reflect the impact of quarantine measures. The latter may be fitted by the quarantine measures function *p*(*l*) = 2.12591 10^5^/*l*^4^ (hyperbolic part of curve 2). After this, a “second wave” is obvious. However in this work we restrict ourselves to considering the first wave of the epidemic in order to show the effectiveness of the proposed model.

**Figure 2:**
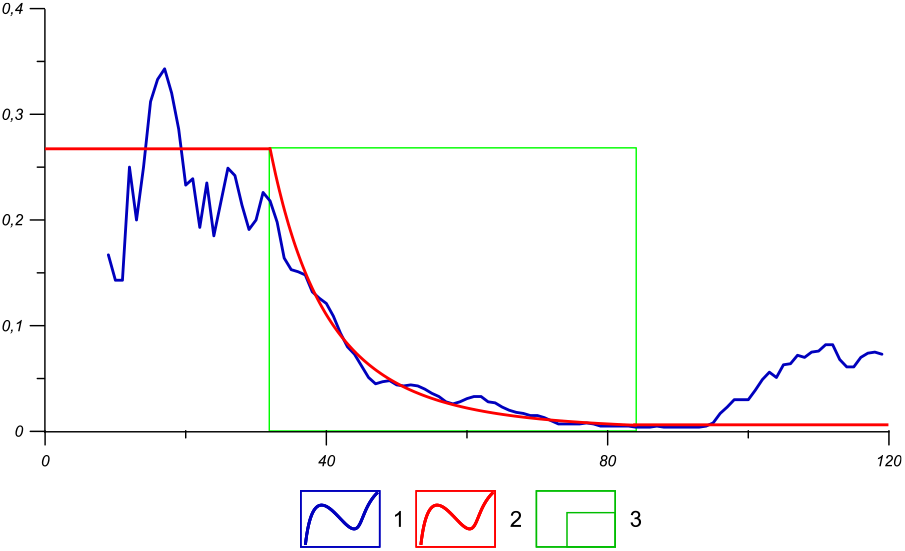
Function *p*(*l*) according to the official data on Israel (population *N* = 8000000, *d* = 16 days) till the day 120 of epidemic. x-axis - days after the first infection, y-axis - *p*(*l*). 1– raw observed data; 2 – modeling curve; 3 - time span, where the quarantine measures with *p*(*l*) are fulfilled.

Figure 3 demonstrates the comparison of the active cases of real statistics for Israel [13] with calculations based on Eq. (15) for the fixed value of the average disease duration *d*. The function *n*_*l*_ has been found on the basis of the real data for *N*_*I*_ (*l*) by inverse calculation. By reverse calculations for real statistical data the function *n*_*l*_ has been found. As we see *d* = 16 days seems a suitable value of average *d* to describe the curve of real active cases of COVID-19 in Israel. This value of *d* is in a reasonable agreement with the statistical average value of the disease duration. However, the average duration of disease *d* can be different for different countries, populations and even stages of the epidemic.

**Figure 3:**
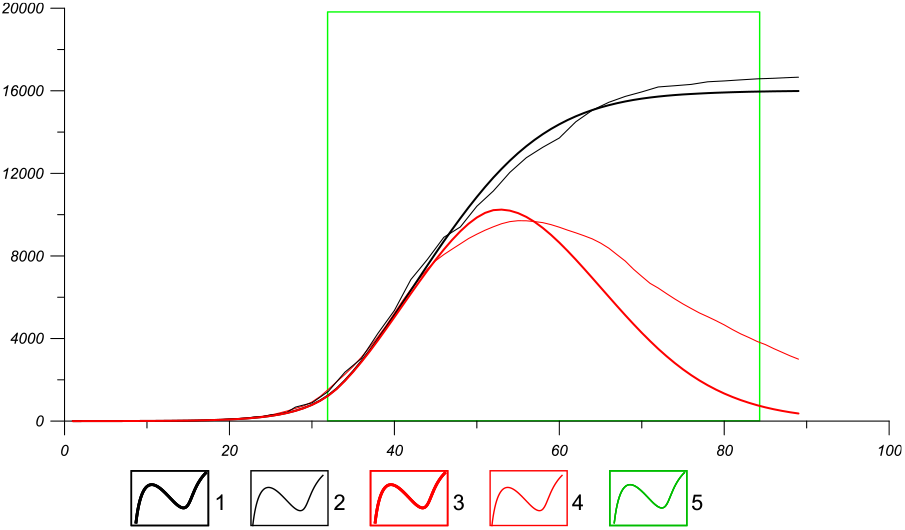
Comparison of computed and statistical epidemic-data for Israel. x-axis - days after the first infection, y-axis - cases. The data are the same as for figure 2.1– the total cases *N*_*T*_ (*l*) computed; 2 – the total cases, observed; 3 – the active cases (*N*_*I*_ (*l*)) computed; 4 – the active cases observed; 5 – the hyperbolic stage from 34 to 83 days.

The similar calculations for Switzerland with appropriate fitting parameters are presented on figures 4 and 5. The hyperbolic approximation *Const*/*l*^4^ as for Israel is a good approximation for the period of the quarantine measures. However the value of constant is different. The initial conditions for the performed calculations corresponds for both countries the real statistical data [19].

**Figure 4:**
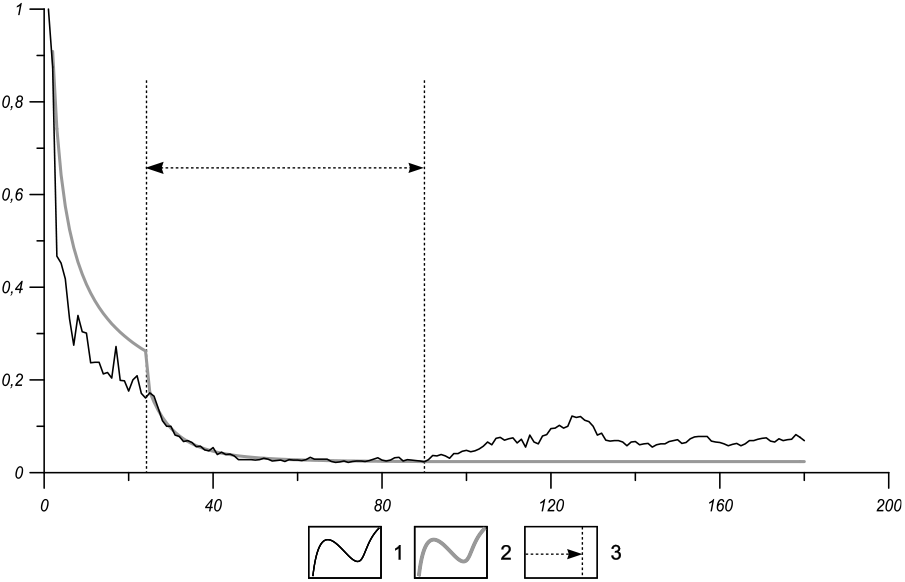
Function *p*(*l*) according to official data on Switzerland (population *N* = 8800000, *d* = 18 days) till the day 180 of epidemic. x-axis - days after the first infection, y-axis *p*(*l*). 1 –observed data with 7-days running average; 2 – modeling curve; 3 - time span from 25 to 90 days, where the quarantine measures are fulfilled resulting in a hyperbolic decrease of *p*(*l*) (*p*(*l*) = 0.023 + 59124*l*^−4^). In the initial stage of the epidemic(*l* < 25) *p* = 0.35. At the post-quarantine stage (*l* > 90) *p*(*l*) = *p*(90) = 0.025.

**Figure 5:**
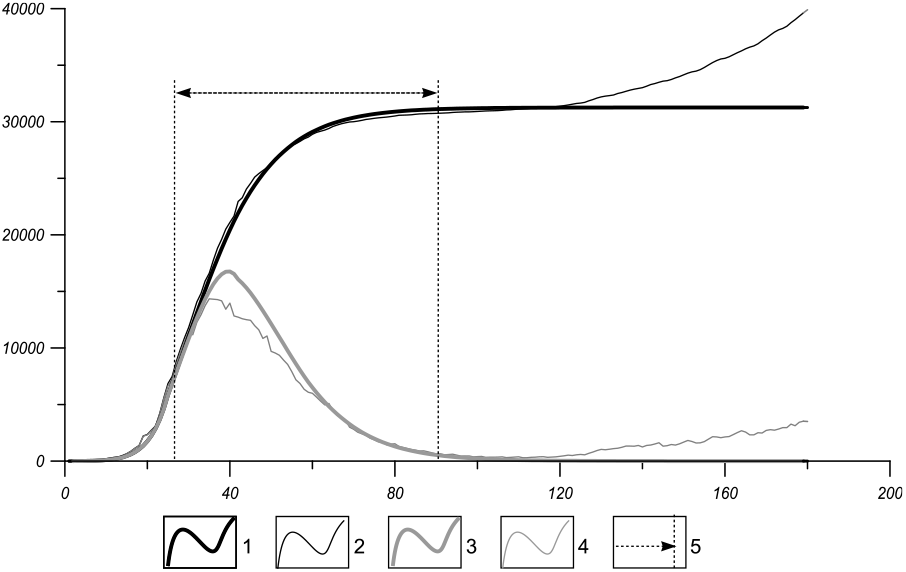
Comparison of computed and statistical epidemic-data for Switzerland. x-axis - days past the first infection; y cases. The data are the same as for figure 4. 1– the total cases *N*_*T*_ (*l*) computed; 2 – the total cases, observed; 3 – the active cases (*N*_*I*_ (*l*)) computed; 4 – the active cases observed; 5 – the hyperbolic stage from 25 to 90 days.

### IV. DELAY DIFFERENTIAL EQUATION

Instead of using discrete time, *l*, in Eq. (7) one can come to continuous time description. To do so we denote *t* = Δ*t*(*l* − 1), where Δ*t* is a unit of time that equals to one day, and put *N*_*T*_ (*l*) = *x*(*t*)*N, N*_*T*_ (*l*) *N*_*T*_ (*l* − 1) = *x*′(*t*)*N*. Then Eq. (7) is rewritten as

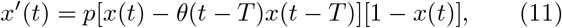

where *θ*(*t*) is the Heaviside’s step function and *T* = *d* − 1. We assume here that time is still measured in days, i.e., Δ*t* = 1, otherwise coefficient *p* should be renormalized. At the initial stage of an epidemic, 0 ≤ *t* ≤*T*, Eq. (11) reads

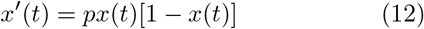

and solution to it is given by

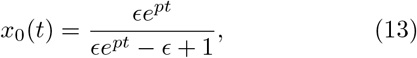

where *x*(0) =, *ϵ*. Solution (13) serves as an initial history function for Eq. (11), that is, *x*(*t*) = *x*_0_(*t*) at 0 ≤ *t* ≤ *T*. There are two stationary solutions to Eq. (12), namely, the unstable solution *x*_0_(*t*) = 0 and the stable solution *x*_0_(*t*) = 1 which is the limit of *x*_0_(*t*) (10) at *t* → ∞. In contrast, delay differential equation (11) has an arbitrary stationary solution *x*(*t*) = *C* at *t* > *T* (0 ≤ *C* ≤1). Evidently, at large time any solution to Eq. (11) tends to some stationary stable saturation value as depicted in figure 1.

The minimal saturation value may be estimated by linearizing Eq. (11) near an arbitrary stationary solution *x*(*t*) = *C*. Let *x*(*t*) = *C* + *δx*(*t*) then linearized equation is *δx*′ (*t*) = *p*(1 − *C*)[*δx*(*t*) − *δx*(*t* − *T*)]. Assuming that *δx*(*t*) = exp(*λt*/*T*) we arrive at the characteristic equation

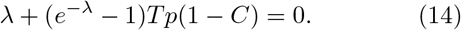

One of the roots of this equation is *λ* = 0. Another real root of Eq. (14) is negative *λ* < 0 if *Tp*(1 − *C*) < 1. In other words, stationary solution *x*(*t*) = *C* is stable if

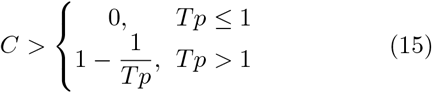

Therefore, the saturation value *x*_∞_(*p*) = lim_*t*→∞_ *x*(*t*) should exceed the value given by Eq. (15). Figure 6 shows dependence of asymptotic value of solution to Eq. (11) on *pT* calculated for two values of initial perturbation, *ϵ* = 10^−5^ (curve 1) and *ϵ* = 2 10^−2^ (curve 2). If *pT* < 1, there is no significant growth of epidemics. For larger values of *pT* the saturation level is always larger than the value given by Eq. (15) and tends to unity for *pT* ≫ 1. For small enough value of *ϵ* ≪ 1, the function *x*_∞_(*p*) tends to a universal curve that is independent of initial perturbation (e.g. curve 1 in Fig. 2). However, the time required to reach the saturation value depends on initial perturbation.

**Figure 6:**
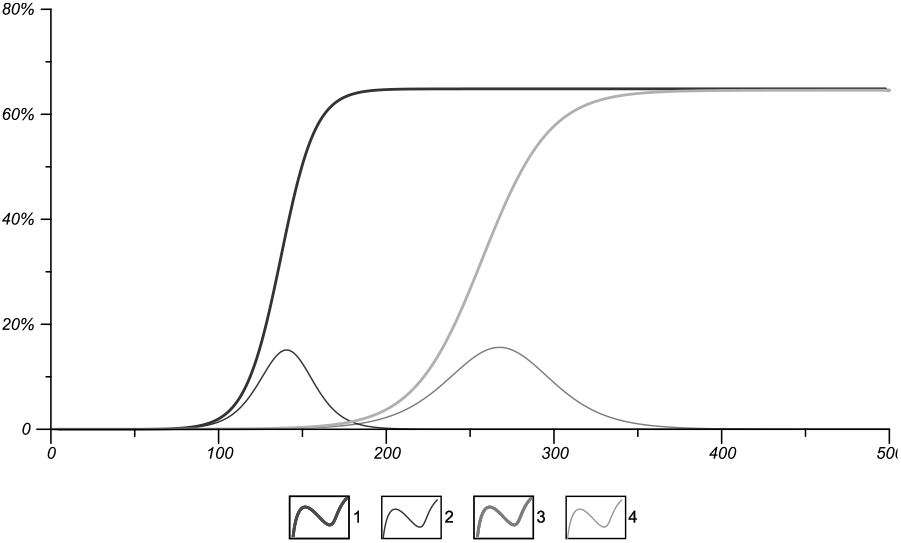
Parametrical dependence of the model under consideration, x-axis - days after first case of infection of the epidemic in a closed population; y-axis - cases of infection as a percent of whole population: 1, 3 - total confirmed cases of infection ; 2, 4 - active cases of infection for a given day; 1, 2 - *p* = 0.16, *d* = 11, *pT* = 1.6; 3, 4 - *p* = 0.08, *d* = 21, *pT* = 1.6.

### V. COMPARISON WITH THE SIR-MODEL

As is known the existing models of epidemic spread are based on two models susceptible-infected-susceptible (SIS) and susceptible - infected - recovered (SIR)(see, e.g., [10, 11]). In the first model the recovered people are immediately transfer to the group of susceptible and can be again infected. In the second model the recovered people save immunity. Therefore, the SIR model is more close to the above considered model and we have compare these models. The main equations of the SIR model (the particular case of the Lotka-Volterra equations [27, 28]) for the necessary values of total and active cases may be modified as

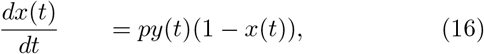

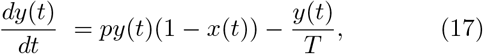

where *x*(*t*) = *N*_*T*_ (*t*)/*N* and *y*(*t*) = *N*_*I*_ (*t*)/*N* correspond to the fractions of population that are affected and infected, respectively.

Taking into account the first integral of set (16), (17) *y*(*t*) = *ϵ* + *x*(*t*) + ln(1 − *x*(*t*))/(*pT*), where it assumed that initially *x*(0) = 0 and *y*(0) = *ϵ*, we arrive at single first-order equation

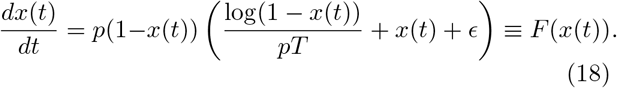

The examples of epidemic growth calculated using two models (11), (18) are depicted in fig. 8. For small enough initial perturbation, *ϵ*, the SIR model predicts approximately twice longer time of saturation. As it was already mentioned there are an infinite number of stationary states of delay equation (11). In contrast, there is only one root of the equation *F* (*x*_1_) = 0 that may be written as *x*_1_(*ϵ, pT*) = 1 + *W* (− *pTe*^−*pT* (*ϵ*+1)^) /(*pT*), where *W* (*z*) is the Lambert function (e.g. [29]). It is easy to verify that the stationary state is stable, i.e., *F*′ (*x*_1_) > 0. It was numerically found that for sufficiently small value of initial perturbation, *ϵ* ≪ 1, the saturation level given by both models are nearly equal. For example, if *ϵ* = 10^−5^ then the dependence *x*_1_(*ϵ, pT*) on *pT* practically coincides with curve 1 in fig. 6.

**Figure 7:**
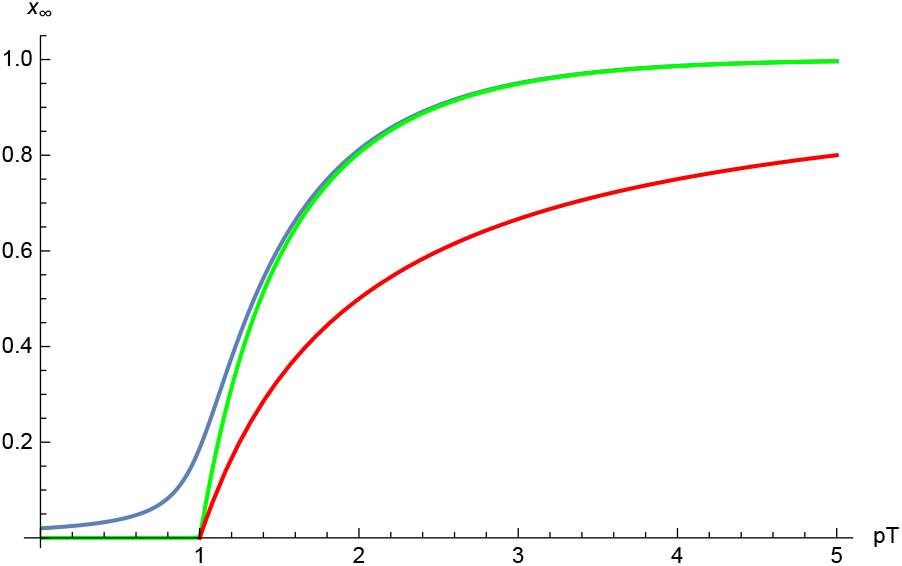
Dependence of saturation value *x*_∞_ on *pT*. Curve 1 — *ϵ* = 10^−5^, curve 2 — *ϵ* = 210^−2^, curve 3 is given by Eq. (15).

**Figure 8:**
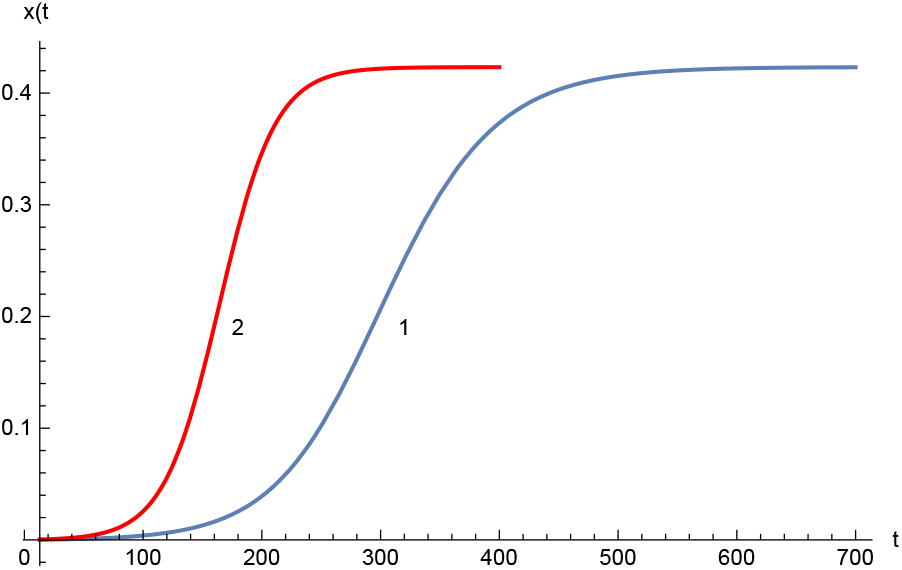
Comparison of two models. Curve 1 — SIR model (18), curve 2 - delay equation (11). = 10^−4^, *p* = 0.1, *T* = 13

## VI. CONCLUSIONS

The real-world epidemic course requires discrete approach to the description which can be useful for determination of the strategy of the quarantine measures. Instead the SIR-type model, based on the differential equations, we derive the non-differential equations in discrete time (days). This approach corresponds to the real daily statistical reports about epidemic course. The transferring from discrete time model to differential equation shows the essential difference in comparison with the SIR model due to the delay obliged by the long duration of COVID-19.

The essential novelty of the obtained results is also the consequent inclusion of quarantine measures in general form through the time-dependent “function of an external influence” *n*_*l*_ (*p*(*l*)). The role of the different quarantine measures and numerical estimation of their influence is still not clear and discussed. Nevertheless, the model under consideration permits to find this function by the reverse calculation on the basis of existent discrete statistical data. The results can be used to assess the impact of quarantine measures for selected regions and countries in general, since the dates and types of regulations are known. This is a way to understand the effectiveness of the regulation rules. We used the reverse calculations to reconcile the values of the theory parameters with observational data. It is possible by use enough simple phenomenological approach. We suppose that this phenomenological approach can be useful for different regions and countries, but cannot be universal due to various regulations, related with economics, national traditions and level of subordination of people to the suggested quarantine measures [19, 20]. Not only constants in the function of external influence *p*(*l*) ≃ *a* + *b*/*l*^4^, as for the case of Germany and Israel, but also the functional dependence *p*(*l*) on *l* can be different for different countries. We also show and investigate the appearance of the collective immunity under quarantine measures. It is the property of the considered deterministic (and in this sense non-Markovian) model with the delayed process of transferring from the subset of virus carriers *V* to the subset of recovered people *R*, which is absent in the known SIR and SEIR models and their modifications. This delay is the characteristic peculiarity of the COVID-19. We demonstrate, that this type of the collective immunity, depended on the parameters of infectivity *p*(*l*) (or *p*(*t*) in differential form with continues time), average duration *d* of the virus transmission time (active cases) and the initially infected part of population ϵ, can be supported on a low level. We have mentioned the models that take into account the change in the infectiousness of the patient in the course of his illness. They are suitable for those cases when the course of the epidemic in small populations is observed in full detail [30, 31]. However, for large populations, such details cannot be known, so their modeling cannot be justified and useful. For such populations, information about the quarantine conditions in which the epidemic develops is much more important. The qualitative confirmation of the obtained in the framework of the considering model results demonstrates the experience of China ans S. Korea. This conclusion supports the use of reasonable strong (taking into account, however, the economical consequences and population fatigue) quarantine measures till the wide vaccination process. Moreover, the model permits to calculate the necessary level of vaccination to reach collective immunity under vaccination. However, these calculations are outside the purposes of this article.

In the suggested model we considered the free epidemic process for various constant values of the average dangerous contacts *n*_*c*_ [22, 23] of one infected individual per day. The idealistic equation for infinite population is generalized for finite population. The general equations are derived for finite population for both cases - with and without quarantine measures. The parameters for saturation of epidemic on the level of total cases *N*_*T*_ < *N* are determined. The developed theory permits to predict the optimal quarantine measures. On the example of Israel and Switzerland a good agreement between the considered theoretical model and the statistical data is found.

We restrict our consideration of epidemic in Israel by the period before the “second wave” (in sense of new essential increase of the active cases) started recently. The second wave for Switzerland also exists but is less pronounced. However, the presented theory can describe all the process, including new waves of COVID-19 observing in present and periodical change of the balance between quarantine measures and reduction of the quarantine restrictions.

## Data Availability

All reffered in the manuscript data are free available

https://www.worldometers.info/coronavirus

## Acknowledgment

The authors are thankful to G.J.F. van Heijst, I.M. Sokolov and A.G. Zagorodny for the useful discussions. S.T. thanks L. Fedulova and M. Karavaeva for stimulating interest to this work.

## Notes

### Competing Interest Statement

The authors have declared no competing interest.

### Funding Statement

No funding for this research

### Author Declarations

I confirm that the submitted work did not require ethical oversight by a dedicated ethics oversight body. All ethical guidlines are completely fulfilled.

## References

[1] Kermack, W.O., and McKendrick, A.G., A contribution to the mathematical theory of epidemics, Proc. Royal Soc. A 115, 700 – 721, https://doi.org/10.1098/rspa.1927.0118 (1927)

[2] Brauer F.; Castillo-Chavez C., Mathematical Models in Population Biology and Epidemiology. Springer-Verlag, 2000

[3] Ball, F., Stochastic and deterministic models for SIS epidemics among a population partitioned into households, Math. Biosci., 156, 41 (1999)

[4] Keeling, M., and Eames, K., Networks and epidemic models, J.R. Soc. Interface 2, 295–307 (2005)

[5] Chowell, G., Sattenspiel, L., Bansald, S., and Viboud, C., Mathematical models to characterize early epidemic growth: A review, Physics of Life Reviews 18, p. 66–97 (2016)

[6] Bedford J., Farrar J., Ihekweazu C., Kang G., Koopmans M. and Nkengasong J., A new twenty-first century science for effective epidemic response, Nature 575, p. 130–136 (2019)

[7] Ghoshal, G., Sander, L.M. and Sokolov, I.M., SIS epidemics with household structure: the self-consistent field method, arXiv:cond-mat/0304301 v1 [cond-mat.stat-mech] 12 Apr 2003

[8] Frey, F., Ziebert, F., and Schwarz, U., Stochastic Dynamics of Nanoparticle and Virus Uptake, Phys. Rev. Lett. 122, 088102 (2019)

[9] Nakamura, G.M., and Martinez, A.S., Hamiltonian dynamics of the SIS epidemic model with stochastic fluctuations, Scientific Reports 9, 15841 (2019)

[10] Sander L.M., C. P. Warren C.M., Sokolov I.M., Epidemics, disorder, and percolation Physica a: Statistical Mechanics and Its Applications, 325, 1 (2003)

[11] Murray J.D., Mathematical Biology, Springer Verlag, New York, 1993.

[12] Kevin Heng, Christian L. Althaus, The approximately universal shapes of epidemic curves in the Susceptible Expo sed Infectious Recovered (SEIR) model, Scientific Reports, 0:19365 (2020)

[13] Xinzhi Liu, Peter Stechlinski, Infectious disease models with time-varying parameters and general nonlinear incidence rate, Applied Mathematical Modelling, V. 36, Issue 5, 1974 (2012)

[14] Marek Kochanczyk, Frederic Grabowski Tomasz Lip- niacki, Dynamics of COVID-19 pandemic at constant and time-dependent contact rates, Math. Model. Nat. Phenom. 15, 28 (2020)

[15] Brahim Boukanjime, Tomas Caraballo, Mohamed El Fatini, Mohamed El Khalifi, Dynamics of a stochastic coronavirus (COVID-19) epidemic model with Markovian switching, Chaos, Solitons and Fractals 141, 110361 (2020)

[16] Xuerong Mao, Glenn Marion, Eric Renshaw, Environmental Brownian noise suppresses explosions in population dynamics, Stochastic Processes and their Applications 97, 95, (2002)

[17] Shaobo He, Yuexi Peng, Kehui Sun, SEIR modeling of the COVID-19 and its dynamics, Nonlinear Dynamics, 101, 1667 (2020)

[18] Bommer, C., and Vollmer, S., Press release: COVID-19: on average only 6 prcent of actual SARS-CoV-2 infections detected worldwide, No. 48 - 06.04.2020, www.uni-goettingen.de/vollmer (2020)

[19] Worldometer counter, https://www.worldometers.info/coronavirus/countries (2020).

[20] Drosten, C. https://www.archyworldys.com/german-professor-called-useless-tests-for-antibodies-to-coronavirus (2020)

[21] CNRS Report (France), COVID-19: Une modelisation indique que pres de 6 percent des Francais ont ete infectes, http://www.cnrs.fr/fr/covid-19-une-modelisation-indique-que-pres-de-6-desfrancais-ont-ete-infectes; https://hal-pasteur.archives-ouvertes.fr/pasteur-02548181 (2020)

[22] Trigger S.A., Initial stage of the COVID-19 infection process in human population 2, MedRxiv, https://doi.org/10.1101/2020.04.13.20063701 (2020)

[23] Trigger S.A., Czerniawski E.B., Equation for epidemic spread with the quarantine measures: application to COVID-19, Physica Scripta 95, 105001 (2020).

[24] Trigger S.A., Czerniawski E.B., Ignatov A.M., The model for epidemic transmission with quarantine measures: application to COVID-19, Research Gate, July 2020.

[25] Lavezzo et al., Suppression of a SARS-CoV-2 outbreak in the Italian municipality of Vo’, Nature, 584, 429 (2020)

[26] Hu, B., Guo, H., Zhou, P. et al.. Characteristics of SARS-CoV-2 and COVID-19. Nature Reviews Microbiology (2020)

[27] Lotka A. J., Elements of Physical Biology. Williams and Wilkins (1925)

[28] Volterra V., Variazioni e fluttuazioni del numero d’individui in specie animali conviventi, Mem. Acad. Lincei Roma. 2: 31–113 (1926).

[29] Weisstein E. W. Lambert W-Function. https://mathworld.wolfram.com/LambertW-Function.html

[30] Ganyani et al., Estimating the generation interval for coronavirus disease (COVID-19) based on symptom onset data, Eurosurveillance 25, no. 17, 30 (2020)

[31] Wong et al., Modeling COVID-19 Dynamics in Illinois under Nonpharmaceutical Interventions, PRX 10, 041033 (2020)

